# Validation of the QAMAI tool to assess the quality of health information provided by AI

**DOI:** 10.1101/2024.01.25.24301774

**Authors:** Luigi Angelo Vaira, Jerome R. Lechien, Vincenzo Abbate, Fabiana Allevi, Giovanni Audino, Giada Anna Beltramini, Michela Bergonzani, Paolo Boscolo-Rizzo, Gianluigi Califano, Giovanni Cammaroto, Carlos M. Chiesa-Estomba, Umberto Committeri, Salvatore Crimi, Nicholas R. Curran, Francesco di Bello, Arianna di Stadio, Andrea Frosolini, Guido Gabriele, Isabelle M. Gengler, Fabio Lonardi, Antonino Maniaci, Fabio Maglitto, Miguel Mayo-Yáñez, Marzia Petrocelli, Resi Pucci, Alberto Maria Saibene, Gianmarco Saponaro, Alessandro Tel, Franco Trabalzini, Eleonora M.C. Trecca, Valentino Vellone, Giovanni Salzano, Giacomo De Riu

## Abstract

**Objective:** To propose and validate the Quality Assessment of Medical Artificial Intelligence (QAMAI), a tool specifically designed to assess the quality of health information provided by AI platforms.

**Study design:** observational and valuative study

**Setting:** 27 surgeons from 25 academic centers worldwide.

**Methods:** The QAMAI tool has been developed by a panel of experts following guidelines for the development of new questionnaires. A total of 30 responses from ChatGPT4, addressing patient queries, theoretical questions, and clinical head and neck surgery scenarios were assessed. Construct validity, internal consistency, inter-rater and test-retest reliability were assessed to validate the tool.

**Results:** The validation was conducted on the basis of 792 assessments for the 30 responses given by ChatGPT4. The results of the exploratory factor analysis revealed a unidimensional structure of the QAMAI with a single factor comprising all the items that explained 51.1% of the variance with factor loadings ranging from 0.449 to 0.856. Overall internal consistency was high (Cronbach’s alpha=0.837). The Interclass Correlation Coefficient was 0.983 (95%CI 0.973-0.991; F(29,542)=68.3; *p*<0.001), indicating excellent reliability. Test-retest reliability analysis revealed a moderate-to-strong correlation with a Pearson’s coefficient of 0.876 (95%CI 0.859-0.891; *p*<0.001)

**Conclusions:** The QAMAI tool demonstrated significant reliability and validity in assessing the quality of health information provided by AI platforms. Such a tool might become particularly important/useful for physicians as patients increasingly seek medical information on AI platforms.

## INTRODUCTION

Artificial Intelligence (AI) has brought about a sea change in numerous fields, with healthcare standing out as one of the most significantly impacted [1,2]. Among the myriad AI models available, OpenAI’s (San Francisco, CA, USA) Chat-based Generative Pre-trained Transformer (ChatGPT) has been particularly striking in its reach and influence in just a few months [3,4].

In healthcare, potential applications of ChatGPT could extend well beyond simply providing health-related information to patients and decision-support for healthcare professionals [5–7]. For patients, it could serve as a tool for health education, possibly aiding in understanding symptoms, deciphering diagnoses, and adhering to treatment plans more effectively [8–10].

Regarding healthcare professionals, the capabilities of ChatGPT may not be confined to simplifying complex medical terminologies or offering diagnostic assistance. It’s has also been proposed for tasks such as reviewing and summarizing extensive medical literature [11], assisting in the design and implementation of research studies [12], proofreading scientific articles [13], and even potentially aiding in the education of medical students [14]. These findings indicate that by enabling real-time, accessible, and personalized information, ChatGPT could potentially help evolve healthcare delivery into a service that is more effective, efficient, and patient-friendly [15]. It is essential, however, to recognize that while these possibilities are exciting, they are still largely in the realm of potential and have yet to be fully studied or validated.

Despite its potential benefits, the use of AI platforms like ChatGPT in healthcare also presents significant risks that must be thoroughly addressed. First and foremost, inaccuracies or misinterpretations of medical information can lead to dangerous health consequences [15,16]. Additionally, there is the risk of over-reliance on AI for health information and decision-making. Patients may take the information provided by the AI as absolute truth, potentially leading to neglect of professional medical advice. Similarly, medical professionals must not abdicate their clinical judgment and decision-making responsibilities to AI, despite its capacity to process and analyze vast amounts of data rapidly [17]. Privacy and data security concerns are another risk associated with the use of AI in healthcare [18].

Moreover, there are ethical implications to consider, such as ensuring that the use of AI platforms does not contribute to health inequities. There is a risk that those who cannot access or use these technologies due to digital illiteracy, lack of internet access, or other barriers may be left behind [19]. Lastly, the regulatory landscape for AI in healthcare is complex and still evolving. There are challenges associated with ensuring that AI models like ChatGPT comply with regulations related to accuracy, transparency, accountability, and patient safety [20].

Given these risks, rigorous, ongoing evaluation of the quality of health information provided by AI platforms is critical.

Despite the existence of several tools to assess the quality of online health information [21–23], these do not translate effectively to AI platforms. These tools have primarily been designed for manual, human-centric assessments and are not compatible with AI-generated outputs. To date, no validated tool exists to accurately assess the health information provided by ChatGPT, and the few clinical studies published on this topic have used non-validated instruments [9,10,17,24–27].

Recognizing these limitations, and acknowledging the critical gap that exists in the assessment of information quality from AI platforms like ChatGPT, this study aims to bridge this lacuna. We propose and validate the Quality Analysis of Medical AI (QAMAI), a novel tool designed specifically to assess the quality of health information offered by AI platforms regarding otorhinolaryngology, head and neck surgery.

## MATERIALS AND METHODS

### Working group

In February 2023, an international collaborative group was established, composed of maxillofacial surgeons, otorhinolaryngologists, and head and neck surgeons from the Italian Society of Maxillofacial Surgery and the young members’ section of the International Federation of Otorhinolaryngology Societies. The group included researchers from different centers around the world, with the aim of studying the reliability and safety of using AI platforms within the field of head and neck surgery for education, diagnosis, therapy, patient communication, and information processes. For this study, 27 researchers from 25 centers across 5 countries (Italy, Belgium, France, Spain, and the United States) were involved.

The execution of this study did not require the approval of an ethics committee as it did not involve patients or animals. The study was conducted in accordance with the Helsinki principles.

### Quality Analysis of Medical Artificial Intelligence tool development

The QAMAI tool has been developed based on the Modified DISCERN (mDISCERN) instrument [28,29]. The mDISCERN is a well-validated and widely used tool for assessing the quality of health information conveyed by websites [30], social networks [31], YouTube and other multimedia platforms [32]. However, the use of mDISCERN for evaluating information provided by artificial intelligence is not possible as the tool takes into account certain human characteristics such as board certification and the reputation of the content creator, which cannot be applied to artificial intelligence.

The draft of QAMAI was drawn up in English by a group of experts consisting of a public health researcher, two head and neck surgeons, a computer engineer specializing in AI, a bioethics expert, a communications engineer specializing in health communication, a representative of patient associations, and a native English-speaking linguist. The diverse backgrounds of this expert group ensured that the development of QAMAI was comprehensive, and its applicability in diverse contexts was taken into account. In analogy with the mDISCERN, the consensus of experts decided to elaborate a unidimensional construct of the instrument with 6 items, evaluated using Likert scales. The six domains of the information quality were hypothesized to be correlated, just as in mDISCERN, to one dimension: the quality of content of the information itself. Each parameter was evaluated by a Likert scale from 1 (strongly disagree) to 5 (strongly agree). The score was then summed into an overall score (QAMAI score) that identified the quality of the information. The streamlined structure of the tool was intentionally designed to ensure quick application and promote its widespread use, thereby broadening its potential impact.

The first draft of the tool was preliminarily tested by an international sample of researchers who evaluated a set of responses provided by ChatGPT4. The results were reviewed by the consensus of experts, along with feedback provided by the researchers. Any areas of uncertainty or confusion regarding any item were addressed and corrected until the final version of the tool was developed [Table 1].

**Table 1.**
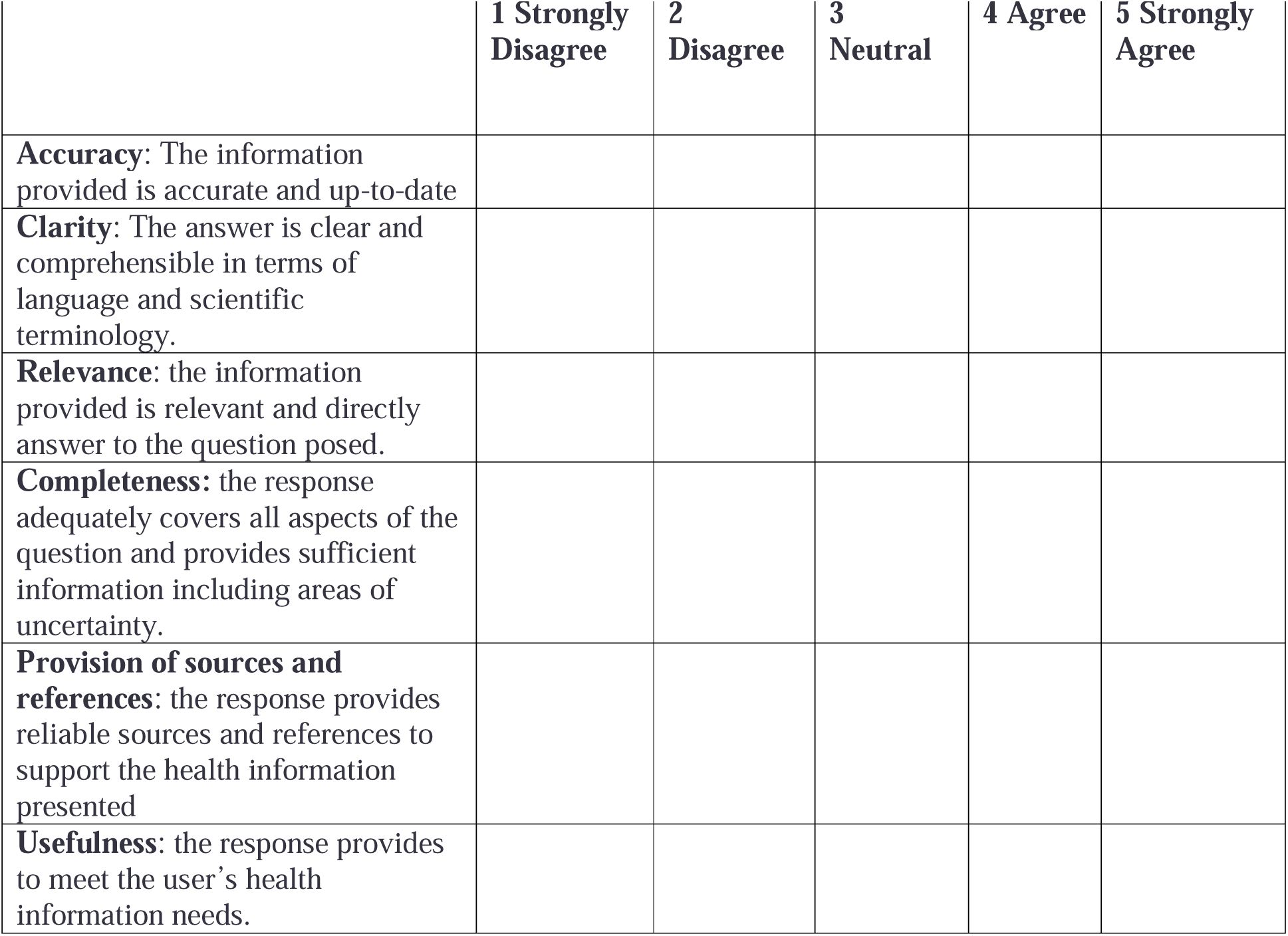
The Quality Analysis of Medical Artificial Intelligence tool.

The QAMAI included six items: accuracy, clarity, relevance, completeness, sources, and usefulness. The QAMAI score, ranging from 6 to 30, allowed the classification of the response into five quality grades [Table 2].

**Table 2.**
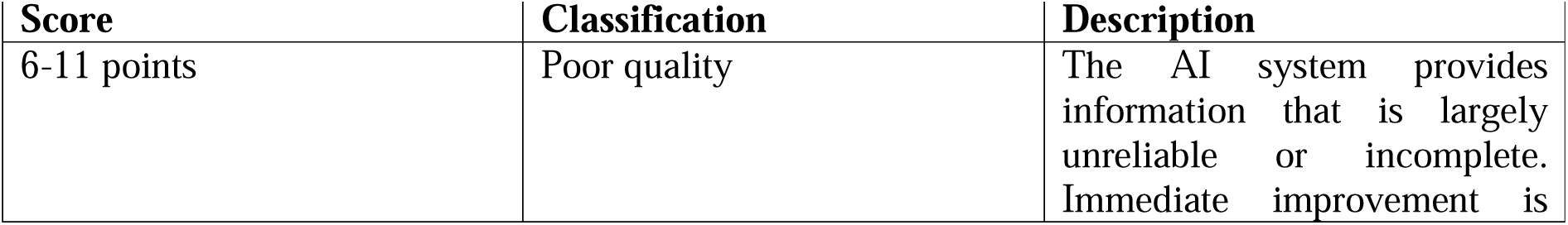
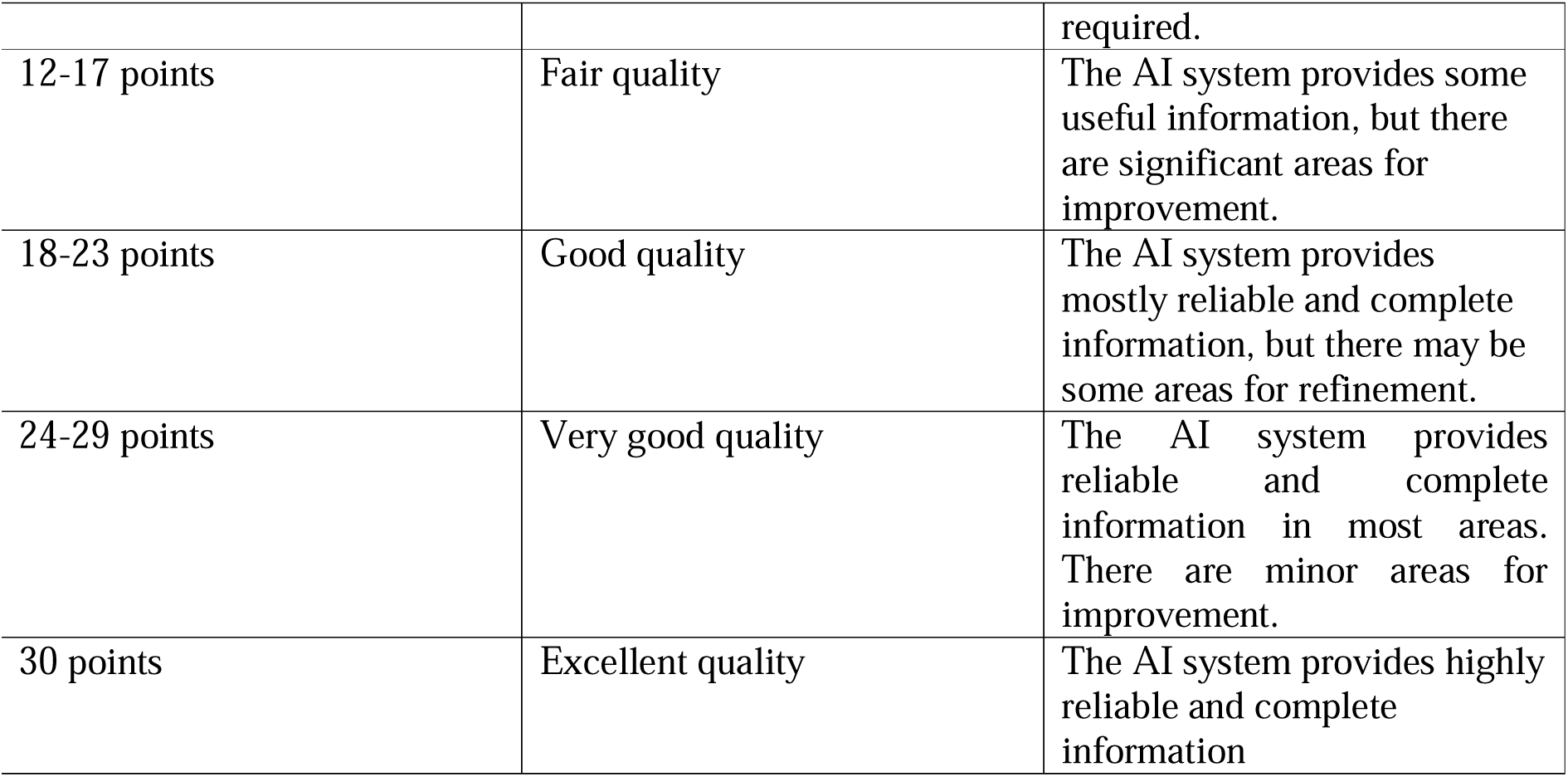
The Quality Analysis of Medical Artificial Intelligence tool scoring system.

### Quality Analysis of Medical Artificial Intelligence tool validation process

A group of three researchers including two head and neck surgeons and a computer engineer specializing in AI prepared a set of 30 questions covering various areas of head and neck surgery. Three types of questions were included: patient inquiries, theoretical questions, and clinical scenarios. The questions were reviewed by the research group and revised if they presented errors or areas of uncertainty until full consensus was reached. The questions were individually entered by a single researcher into the ChatGPT version 4 chatbot on May 25, 2023. The AI was asked to provide the most complete and exhaustive answers possible, including the bibliographic sources from which it drew its information. The exact prompt, used for all the questions, was: “please act as an head and neck surgeon and provide an answer to this question that is as exhaustive and precise as possible, taking into consideration the most recent guidelines and clear scientific evidence you have available and citing the bibliographical sources from which you drew your answers”. The responses were recorded by the researcher for subsequent analysis. The full set of questions and answers is reported in the Supplementary Table 1 [Supplementary Table 1].

The set of answers was provided to a pool of 27 head and neck surgeons specializing in otolaryngology or maxillofacial surgery. The researchers were asked to independently evaluate the responses using the QAMAI tool, refraining from giving an evaluation if the subject matter was beyond their knowledge. The evaluation was repeated a second time, 10 days after the first. The responses obtained from the 27 researchers were collected and analyzed for the validation of the QAMAI tool.

### Statistical analysis

Statistical analyses were performed using Jamovi version 2.3.18.0, a freeware and open statistical software available online at www.jamovi.org [33]. Categorical variables are reported in numerals and percentages of the total. Descriptive statistics for quantitative variables are given as the median (interquartile range (IQR)) or mean ± standard deviation (SD). The QAMAI score differences among the three categories of questions were assessed using a one-way ANOVA; if significant differences were found, Dwass-Steel-Crichlow-Fligner test was employed for post-hoc analysis.

For the validation of the QAMAI, the number of questions and respondents was preliminarily determined with the aim of having at least 30 responses to evaluate for each item of the tool. The resulting sample size should be considered excellent [34]. Furthermore, Bartlett’s test of sphericity and Kaiser-Meyer-Olkin (KMO) test were used to assess the sampling adequacy. A KMO value > 0.6 indicates adequate sampling [35].

For the construct validation of the questionnaire, an exploratory factor analysis was conducted to uncover the inter-relations between clusters of items and the number of factors assessed by the questionnaire. In order to maximize the loading of each variable on the extracted factors a minimum residual extraction method and a promax rotation with a cut-off point of 0.40 and the Kaiser’s criterion of eigenvalues greater than 1 were used for the analysis. Upon identification of the number of factors through exploratory factor analysis, a confirmatory factor analysis was conducted to verify if the data would fit the specific theoretical model that had been identified. The assessment of the model’s goodness-of-fit was carried out using the Root Mean Square Error of Approximation (RMSEA) and the Comparative Fit Index (CFI). Indicators of a good model fit were considered to be RMSEA values of less than 0.05 and CFI values greater than 0.95 [36].

Internal consistency was then assessed using Cronbach’s alpha, to determine whether the tool’s items were inter-correlated and consistent with the tool’s construct. A Cronbach’s alpha of at least 0.70 has been suggested to indicate adequate internal consistency [37].

Inter-rater reliability was assessed by comparing, for each question, the evaluations provided by the different reviewers using intraclass correlation coefficient (ICC). The values considered for ICC were as follows: ICC < 0.5 as poor reliability, ICC = 0.5 − 0.75 as moderate reliability, ICC = 0.75 − 0.9 as good reliability, ICC > 0.9 as excellent reliability [38].

Finally, the test-retest reliability between the two evaluations provided by researchers 10 days apart was assessed using Pearson’s correlation coefficient. For all tests, the level of statistical significance was set at p<0.05.

## RESULTS

Twenty-seven reviewers provided a total of 792 assessments for the 30 responses given by ChatGPT (18 assessments missing in the dataset). The median QAMAI scores reported for each question are displayed in Supplementary Table 1. Out of the 792 quality assessments collected, 30 were found to be excellent (i.e., QAMAI score 30), 261 very good (i.e., QAMAI score 24-29), 399 good (i.e., QAMAI score 18-23), 100 fair (i.e., QAMAI score 12-17), and 3 poor (i.e., QAMAI score 6-12). The differences in QAMAI scores across the three categories of questions (patient questions, theoretical questions, and clinical scenarios) were not found to be statistically significant (one-way ANOVA: χ^2^=3.86; *p*=0.145).

To evaluate the adequacy of the sampling, the Bartlett’s test of sphericity was initially performed, which returned a significant result (χ^2^=2005; *p*<0.001), indicating the appropriateness of conducting the KMO test. This latter test yielded an overall measure of sampling adequacy of 0.882, indicating an excellent sample size.

The results of the exploratory factor analysis revealed a unidimensional structure of the QAMAI with a single factor comprising all the items that explained 51.1% of the variance with factor loadings ranging from 0.449 to 0.856 [Table 3].

**Table 3.** Exploratory factor analysis results.

|  | **Factor** |  |
| --- | --- | --- |
|  | **1** | **Uniqueness** |
| Accuracy | 0.743 | 0.448 |
| Clarity | 0.681 | 0.536 |
| Relevance | 0.772 | 0.403 |
| Completeness | 0.719 | 0.484 |
| Sources | 0.449 | 0.798 |
| Usefulness | 0.856 | 0.267 |

Note. 'Minimum residual' extraction method was used in combination with a 'promax' rotation

The evaluation of the goodness-of-fit confirmed a good fit between the one factor model and the data, revealing an RMSEA value of 0.053 [90% confidence interval 0.033-0.075] and a CFI value of 0.989.

The internal consistency of the questionnaire was evaluated using Cronbach’s alpha, which yielded a value of 0.837 confirming an excellent internal consistency among the items in the questionnaire, suggesting that they measure the same underlying concept or construct [Figure 1]. Excellent inter-rater reliability was found comparing reviewers’ QAMAI scores for each answer. The average ICC was 0.983 with a 95% confidence interval from 0.973 to 0.991 [F(29,542) = 68.3; *p*<0.001].

**Figure 1.**
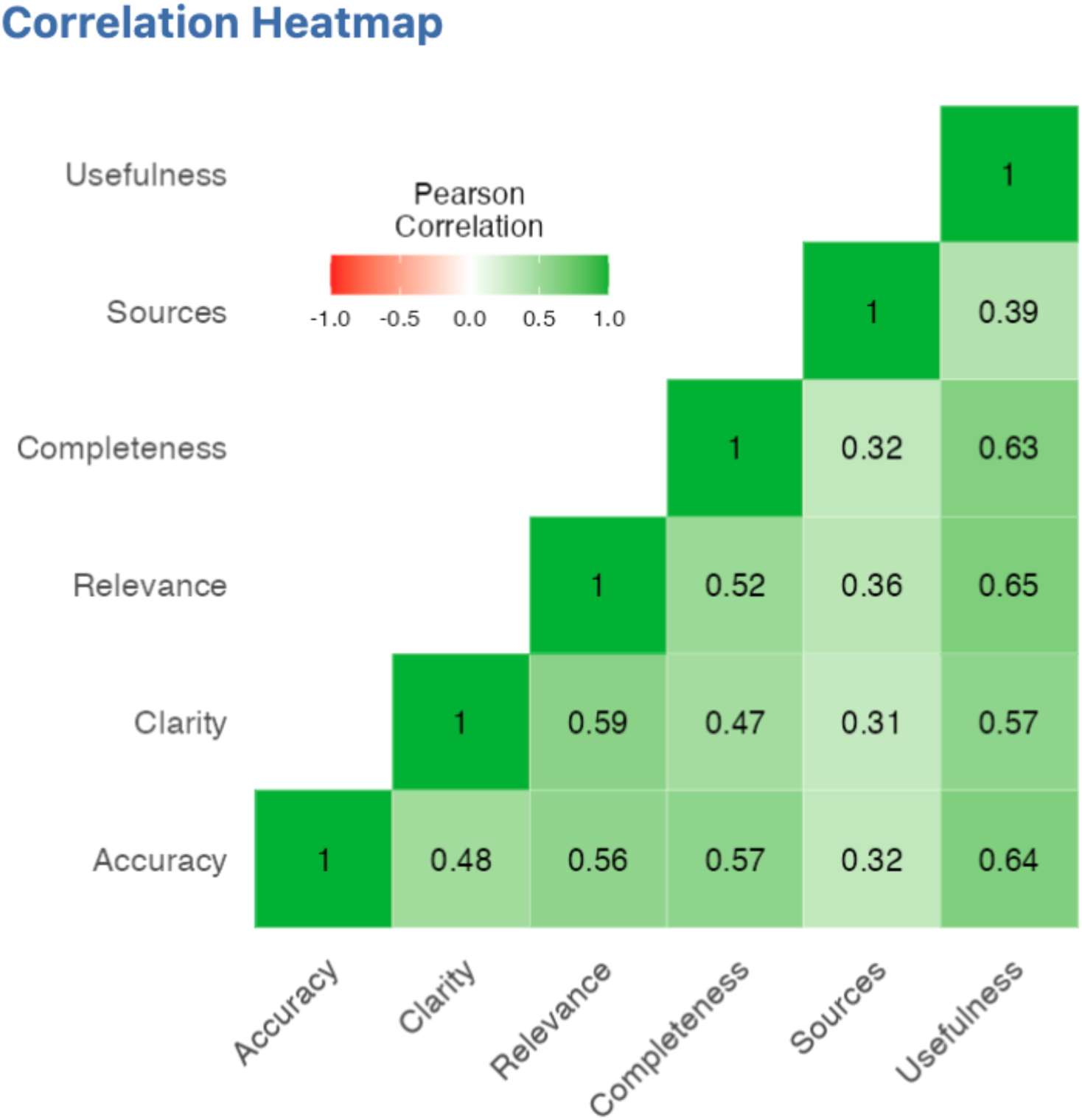
Heatmap of the correlations between QAMAI items.

In order to evaluate the test-retest reliability of the QAMAI tool, a Pearson’s correlation test was performed comparing the scores given at two different times, spaced one week apart. The results showed a Pearson’s correlation coefficient of 0.876 (95% confidence interval 0.859-0.891; *p*<0.001) indicating a moderate-to-strong and significant correlation between the two sets of scores [Figure 2].

**Figure 2.**
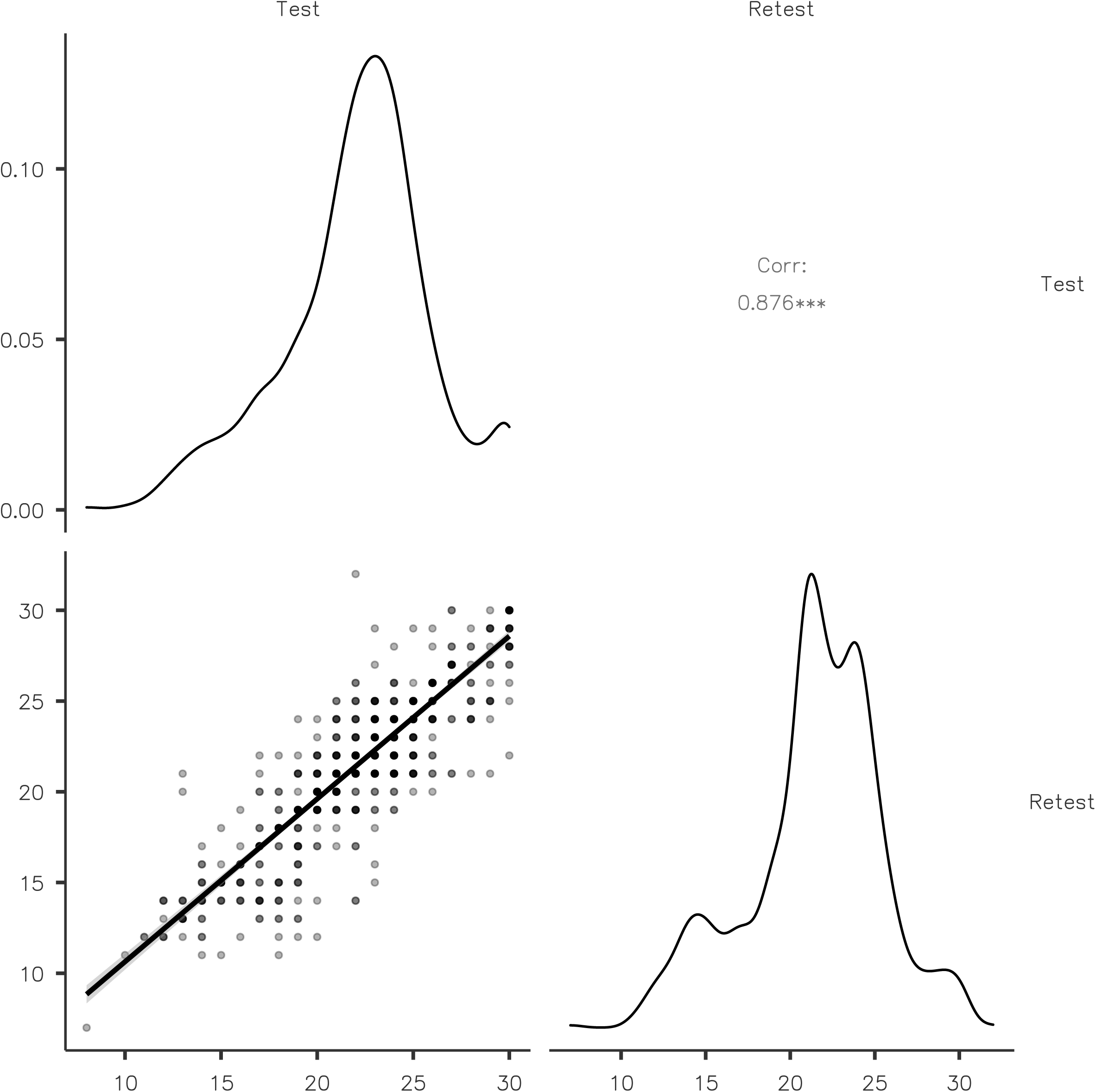
Test-retest reliability analysis.

## DISCUSSION

AI platforms, such as ChatGPT, are poised to revolutionize the way we access and interpret health-related information in the coming years. This innovative application of AI technology offers several potential advantages, but it also presents certain challenges that need to be addressed in the near future. The quality and accuracy of the information provided by these platforms is a significant concern [39,40]. AI platforms rely heavily on the quality and accuracy of the data they have been trained on. Misinformation or outdated data could lead to the provision of incorrect advice, which could have severe consequences in healthcare.

Given these circumstances, it becomes essential to develop and adopt tools to evaluate the quality of health information delivered by AI platforms. These evaluation tools are crucial for several reasons. They first allow for the systematic assessment of AI-generated information’s quality and accuracy, providing an additional layer of scrutiny. Secondly, they can identify areas of weakness or inaccuracy within the AI platform, guiding its further development and improvement. Furthermore, these tools can aid in promoting transparency and trust in AI systems. They can ensure that the information provided meets certain standards, assuaging user concerns about the reliability of AI-generated advice. Finally, they contribute to the broader effort of promoting digital health literacy, empowering users to critically evaluate and make informed decisions about the health information they encounter online.

The QAMAI tool represents the first attempt to provide a validated, widely usable instrument for assessing health information delivered by AI platforms. The design of the QAMAI tool is lean and streamlined, specifically aimed at enabling a swift but comprehensive assessment of AI responses. This agile/specific design is crucial for several reasons. First, it ensures that the evaluation process does not become overly cumbersome or time-consuming, thereby facilitating the tool’s widespread application. Secondly, while being expedient, the QAMAI tool does not compromise on thoroughness. It is designed to deliver a comprehensive evaluation of the AI responses, looking at a range of aspects to determine the quality of the health information provided.

The exploratory factor analysis and internal consistency assessment have revealed that the QAMAI is a unidimensional tool. This unidimensional factor can be identified with the quality of information, and all instrument items are inherently tied to this core construct. On the internal consistency analysis, the item presenting the weakest correlations with others was the quality of sources [Figure 1]. This is likely tied to the propensity of ChatGPT to often provide correct or near correct answers in content, but with non-existent bibliographic references. Literature reports a rate of erroneous sources exceeding 80% for version 3.5 [41,42]. In this data series, 30.6% of the sources were non-existent, indicating an improvement in version 4 of ChatGPT. However, this rate remains unacceptably high, underscoring the need for further improvements in the AI system’s ability to provide accurate and reliable source references.

Inter-rater reliability was found to be excellent, showcasing the consistency of QAMAI scores across different reviewers. This strongly supports the tool’s reliability in different hands and further validates its robustness. Furthermore, test-retest reliability demonstrated a strong and statistically significant correlation between two sets of scores spaced one week apart. This indicates the stability of the QAMAI scores over time, affirming the reliability of the tool in providing consistent evaluations across different time intervals.

This study presents several limitations that should be acknowledged. The dataset of questions used in this research was limited to those pertaining solely to head and neck surgery, which may limit the generalizability and applicability of QAMAI across other medical branches. Secondly, the questionnaire was designed to evaluate solely the quality of the information provided, and not the information transmission modalities, an element that is often crucial in human communication.

In patient interactions, empathy and understanding of patients’ needs are nearly as important as the quality of the information itself. In the case of AI-generated health information, these human elements may not be fully captured or conveyed. Therefore, further refinement and expansion of tools like QAMAI are needed to account for these factors when assessing the quality of health information directed at patients.

## CONCLUSIONS

The findings of this study demonstrate that the QAMAI tool represents a valid and reliable instrument to evaluate the quality of health information delivered by AI platforms, such as ChatGPT. The implementation and large-scale utilization of such tools are critical for monitoring the quality of this rapidly expanding source of health information, currently largely unverified. Ensuring the accuracy and reliability of AI-generated health information is of utmost importance in preventing potential harm to users independently seeking health advice or information. As AI continues to revolutionize the health information landscape, the need for robust quality control tools like QAMAI will only increase. Therefore, our findings pave the way for future research and validation efforts, ultimately contributing to safer and more effective use of AI in health information delivery.

## Data Availability

All data produced in the present study are available upon reasonable request to the authors

## ACNOWLEDGEMENTS

None

## CONFLICT OF INTEREST

None of the authors has a financial interest in any of the products, devices or drugs mentioned in this manuscript.

## ETHICAL APPROVAL

Ethical committee approval was not required for this study as it did not involve any patients.

## FUNDING

none

## AUTHORS CONTRIBUTIONS

Luigi Angelo Vaira: conceptualization of the work, development of the methodology, data curation, writing the original draft, writing the final draft, final approval.

Jerome R. Lechien: development of the methodology, writing the original draft, final approval.

Vincenzo Abbate: data collection, data curation, revision of the original and final draft, final approval.

Fabiana Allevi: data collection, data curation, revision of the original and final draft, final approval.

Giovanni Audino: data collection, data curation, revision of the original and final draft, final approval.

Giada Anna Beltramini: data collection, data curation, revision of the original and final draft, final approval.

Michela Bergonzani: data collection, data curation, revision of the original and final draft, final approval.

Paolo Boscolo-Rizzo: data collection, data curation, revision of the original and final draft, final approval.

Gianluigi Califano: data curation and analysis, revision of the original and final draft, final approval.

Giovanni Cammaroto: data collection, data curation, revision of the original and final draft, final approval.

Carlos Miguel Chiesa-Estomba: provision of study instrumentation, development of the methodology, review of the first and final draft, final approval.

Umberto Committeri: data collection, data curation, revision of the original and final draft, final approval.

Salvatore Crimi: data collection, data curation, revision of the original and final draft, final approval.

Nicholas R. Curran: data collection, data curation, revision of the original and final draft, final approval.

Francesco di Bello: data curation and analysis, literature review, revision of the original and final draft, final approval.

Arianna di Stadio: data collection, data curation, revision of the original and final draft, final approval.

Andrea Frosolini: data collection, data curation, revision of the original and final draft, final approval.

Guido Gabriele: data collection, data curation, revision of the original and final draft, final approval.

Isabelle M. Gengler: data collection, data curation, revision of the original and final draft, final approval.

Fabio Lonardi: data collection, data curation, revision of the original and final draft, final approval.

Fabio Maglitto: data collection, data curation, revision of the original and final draft, final approval.

Miguel Mayo-Yáñez: data collection, data curation, revision of the original and final draft, final approval.

Marzia Petrocelli: data collection, data curation, revision of the original and final draft, final approval.

Resi Pucci: data collection, data curation, revision of the original and final draft, final approval.

Alberto Maria Saibene: data collection, data curation, revision of the original and final draft, final approval.

Gianmarco Saponaro: data collection, data curation, revision of the original and final draft, final approval.

Alessandro Tel: data collection, data curation, revision of the original and final draft, final approval.

Franco Trabalzini: data collection, data curation, revision of the original and final draft, final approval.

Eleonora M.C. Trecca: data collection, data curation, revision of the original and final draft, final approval.

Valentino Vellone: data collection, data curation, revision of the original and final draft, final approval.

Giovanni Salzano: conceptualization of the work, development of the methodology, data curation, writing the original draft, writing the final draft, final approval.

Giacomo De Riu: conceptualization of the work, development of the methodology, data curation, writing the original draft, writing the final draft, final approval.

## SUPPLEMENTARY TABLE LEGEND

Supplementary table 1. Questions and answers given by ChatGPT-4. Incorrect references are shown in bold.

## Notes

### Competing Interest Statement

The authors have declared no competing interest.

### Funding Statement

This study did not receive any funding

